# Comparison of Fine-Scale Malaria Strata Derived from Population Survey Data Collected Using mRDTs, Microscopy and qPCR in South-Eastern Tanzania

**DOI:** 10.1101/2024.06.24.24309395

**Authors:** Issa H. Mshani, Frank M. Jackson, Elihaika G Minja, Said Abbas, Nasoro S. Lilolime, Faraja E. Makala, Alfred B. Lazaro, Idrisa S. Mchola, Linda N. Mukabana, Najat Kahamba, Alex Limwagu, Rukia. M. Njalambaha, Halfan S. Ngowo, Donal Bisanzio, Francesco Baldini, Simon A. Babayan, Fredros Okumu

## Abstract

**Introduction:** Malaria-endemic countries are increasingly adopting data-driven risk stratification, often at district or higher regional levels, to guide their intervention strategies. The data typically comes from population-level surveys collected by rapid diagnostic tests (RDTs), which unfortunately perform poorly in low transmission settings. Here, we conducted a high-resolution survey of *Plasmodium falciparum* prevalence rate (PfPR) in two Tanzanian districts and compared the fine-scale strata obtained using data from RDTs, microscopy and quantitative polymerase chain reaction (qPCR) assays.

**Methods:** A cross-sectional survey was conducted in 35 villages in Ulanga and Kilombero districts, south-eastern Tanzania between 2022 and 2023. We screened 7,628 individuals using RDTs (SD-BIOLINE) and microscopy, with two thirds of the samples further analyzed by qPCR. The data was used to categorize each district and village as having very low (PfPR<1%), low (1%≤PfPR<5%), moderate (5%≤PfPR<30%), or high (PfPR≥30%) parasite prevalence. A generalized linear model was used to analyse infection risk factors. Other metrics, including positive predictive value (PPV), sensitivity, specificity, parasite densities, and Kappa statistics were computed for RDTs or microscopy using qPCR as reference.

**Results:** Significant fine-scale variations in malaria risk were observed within and between districts, with village prevalence ranging from 0% to >50%. Prevalence varied by testing method: Kilombero was low risk by RDTs (PfPR=3%) and microscopy (PfPR=2%) but moderate by qPCR (PfPR=9%); Ulanga was high risk by RDTs (PfPR=39%) and qPCR (PfPR=54%) but moderate by microscopy (PfPR=26%). RDTs and microscopy classified majority of the 35 villages as very low to low risk (18 - 21 villages). In contrast, qPCR classified most villages as moderate to high risk (29 villages). Using qPCR as the reference, PPV for RDTs and microscopy ranged from <20% in very low transmission villages to >80% in moderate to high transmission villages. Sensitivity was 62% for RDTs and 41% for microscopy; specificity was 93% and 96%, respectively. Kappa values were 0.58 for RDTs and 0.42 for microscopy. School-age children (5-15years) had higher malaria prevalence and parasite densities than adults (P<0.001). High-prevalence villages also had higher parasite densities (Spearman r=0.77, P<0.001 for qPCR; r=0.55, P=0.003 for microscopy).

**Conclusion:** This study highlights significant fine-scale variability in malaria risk within and between districts and emphasizes the variable performance of the testing methods when stratifying risk. While RDTs and microscopy were effective in high-transmission areas, they performed poorly in low-transmission settings; and classified most villages as very low or low risk. In contrast, qPCR classified most villages as moderate or high risk. While we cannot conclude on which public health decisions would be subject to change because of these differences, the findings suggest the need for improved testing approaches that are operationally feasible and sufficiently sensitive, to enable precise mapping and effective targeting of malaria in such local contexts. Moreover, public health authorities should recognize the strengths and limitations of their available data when planning local stratification or making decisions.

## Background

Precise mapping of malaria prevalence is crucial for the eventual elimination of the disease from different localities. In line with World Health Organization (WHO) guidelines, National Malaria Control Programs (NMCPs) in Africa are increasingly adopting data-driven stratification of malaria risk, in most cases at either district or higher regional levels [1–3]. These stratifications involve assessing risk levels in geographical areas at the subnational level (e.g. zones, regions, and districts) [2,4,5], and can include fine scale mapping (down to wards and villages levels) as countries progress towards elimination [6–8]. The data for such stratification may come from health facilities, active malaria screening during population surveys, or proxy data sources such as antenatal care clinic visits [6,9,10].

When developing country-level malaria strategies, the prevalence of malaria, representing the proportion of confirmed positive cases of *Plasmodium falciparum* (or other *Plasmodium* sp.) among all individuals tested [11,12], can be classified into various transmission categories. The WHO has previously used the following cutoff points for malaria endemicities: below 1% as very low, 1-10% as low, 10-35% as moderate, and above 35% as high risk malaria stratum [12]. Different NMCPs may adapt these criteria with slight adjustments based on local epidemiological insights. For instance, some countries, including Tanzania and Kenya, have used the parasite prevalence data to categorize their geographic zones as either very low risk (PfPR < 1%), low risk (1% ≤ PfPR < 5%), moderate risk (5% ≤ PfPR < 30%), or high risk strata (PfPR ≥ 30%) [2,13]. Another measure that can be used for generating these strata is the annual parasite incidence (API), which is the number of diagnostically confirmed malaria cases per 1000 individuals per year and is usually obtained from health facilities data [12,14]. API offers precise malaria stratifications, though it fails to account for sub-clinical malaria infections, which can contribute to transmission and impede malaria elimination efforts [15].

National malaria programs usually rely on different actively and passively collected data to inform malaria burden and monitor the effectiveness of control measures [16–18]. For instance, Tanzania employs multiple platforms, including the District Health Information software (DHIS2) populated with data from routine health facility visits, the Malaria Indicator Surveys (MIS) and Tanzania Demographic and Health Surveys (TDHS), which are done every 4-5 years through household surveys, and the school malaria parasite surveillance targeting kids aged 5-16 years during (SMPS) [2,19–21]. A common feature of these established systems is that most rely primarily on rapid diagnostic tests (RDTs) and microscopy [13,22,23], though samples are sometime also preserved for PCR assays.

Microscopy, long used in malaria diagnosis, can quantify parasite loads and identify different *Plasmodium* species, which are essential for precise treatment choices [24,25]. However, its effectiveness depends significantly on the skill and experience of the microscopist, making it unreliable in some contexts, and it can miss a substantial number of true infections due to sub-optimal accuracy [26–28]. In contrast, Rapid Diagnostic Tests (RDTs) offer a consistent and user-friendly option, enabling quick, on-site diagnosis without specialized skills or equipment. RDTs have become widely used in both point-of-care settings and population surveys due to their operational simplicity and cost-effectiveness [29–33]. While the technique enhances access to diagnostics, especially in remote areas, RDTs have lower sensitivity for detecting low-level infections and cannot quantify parasite density or distinguish between *Plasmodium* species. Additionally, current RDTs may detect antigens for over three weeks post-treatment, leading to poor specificity and potential overestimation of malaria cases in high transmission areas [30,33,34].

In contrast, polymerase chain reaction (PCR) assays are known for their high sensitivity and specificity [35]. While conventional PCR assays typically provide qualitative information on malaria infections, quantitative PCR (qPCR) can offer additional quantitative measures of malaria parasite density [36,37]. Unfortunately, the widespread use of PCR assays for population surveys is hindered by cost constraints and the need for specialized expertise and infrastructure for implementation [35,38,39].

The increased focus on evidence-based strategies in malaria control also includes a transition from broad subnational stratifications to more granular, fine-scale approaches [6,10]. However, although current methods like RDTs and microscopy are favored for their operational simplicity, their effectiveness in detailed risk stratification, which are critical for targeting both clinical and sub-clinical infections for malaria elimination, remains poorly understood. Some authors have also suggested that RDTs may have vastly reduced performance in settings where the malaria burden has been significantly reduced [40]. This calls for a rigorous evaluation and comparison of these methods against highly sensitive techniques such as qPCR to refine malaria stratification approaches for malaria elimination. Indeed, available evidence, including data from Kenya and Tanzania, suggest that PCR assays are generally better at pinpointing main malaria hotspots in communities than RDTs and microscopy [41,42]. The study from Tanzania further showed that in subsequent treatment campaigns relying on RDT-based screening, ∼45% of infections remain untreated, even if treatment is offered to all members of households with an infected individual [42]. In the Kenyan study, the authors went further to suggest that since detection of hotspots depends on the sensitivity of diagnostic tools, health authorities working in malaria elimination settings should consider using PCR to guide detection of the residual hotspots, as this provides greatest opportunities to find asymptomatic individuals and sub-patent parasite reservoirs in the communities [41].

All these studies clearly show that while sub-national stratification may be the most effective approach to decide on how to allocate resources, the type of data used for such epidemiological profiling matters significantly; especially when the stratification is done at local-sub-district levels. In places like southeastern Tanzania, which has experienced decades of sustained malaria interventions and progress, and where robust entomological surveillance already exists [43], addition of detailed parasite prevalence data from population-level surveys is required to enable more precise, fine-scale stratifications at both district and sub-district levels.

The aim of this study was therefore to generate a high-resolution population-level survey map of *P. falciparum* prevalence in two districts in south-eastern Tanzania, and compare the fine scale malaria risk strata obtained when using data from different test methods, namely RDTs, microscopy and qPCR. The study also examined the association between population-level parasite prevalence and the parasite densities as determined by both qPCR and microscopy in the different study villages. Lastly, we investigated the malaria detection capacity of RDTs and microscopy across villages with varying parasite prevalences and densities.

## Methods

### Study site

The study was conducted in Morogoro region, in south-eastern Tanzania (Figure 1), in the two districts of Kilombero (population: ∼583,000; 8.2414°S, 36.3349°E; elevation: ∼270m) and Ulanga (population: ∼233,000; 8.9889°S, 36.6133°E; elevation: ∼800m). The average malaria prevalence in the Morogoro region has previously been estimated to exceed 10%, with *P. falciparum* as the dominant malaria species [19,20,44]. The main economic activities for residents include rice farming, sugarcane farming and maize farming, though the area also has other food crops and large commercial tree plantations (teak). The known annual rainfall range is 1200-1400 mm in the lower-lying plains of Kilombero district, and1400-2100 mm in the higher areas in Ulanga district [45]. Approximately 90% of the rainfall occurs during the wet seasons between December to April, with dry seasons typically lasting from June through September [45]. The annual mean daily temperature is around 27°C in the lowlands and approximately 23°C in the highlands. Relative humidity averages from 75% in the lowlands to 80% in the highlands.

**Figure 1:**
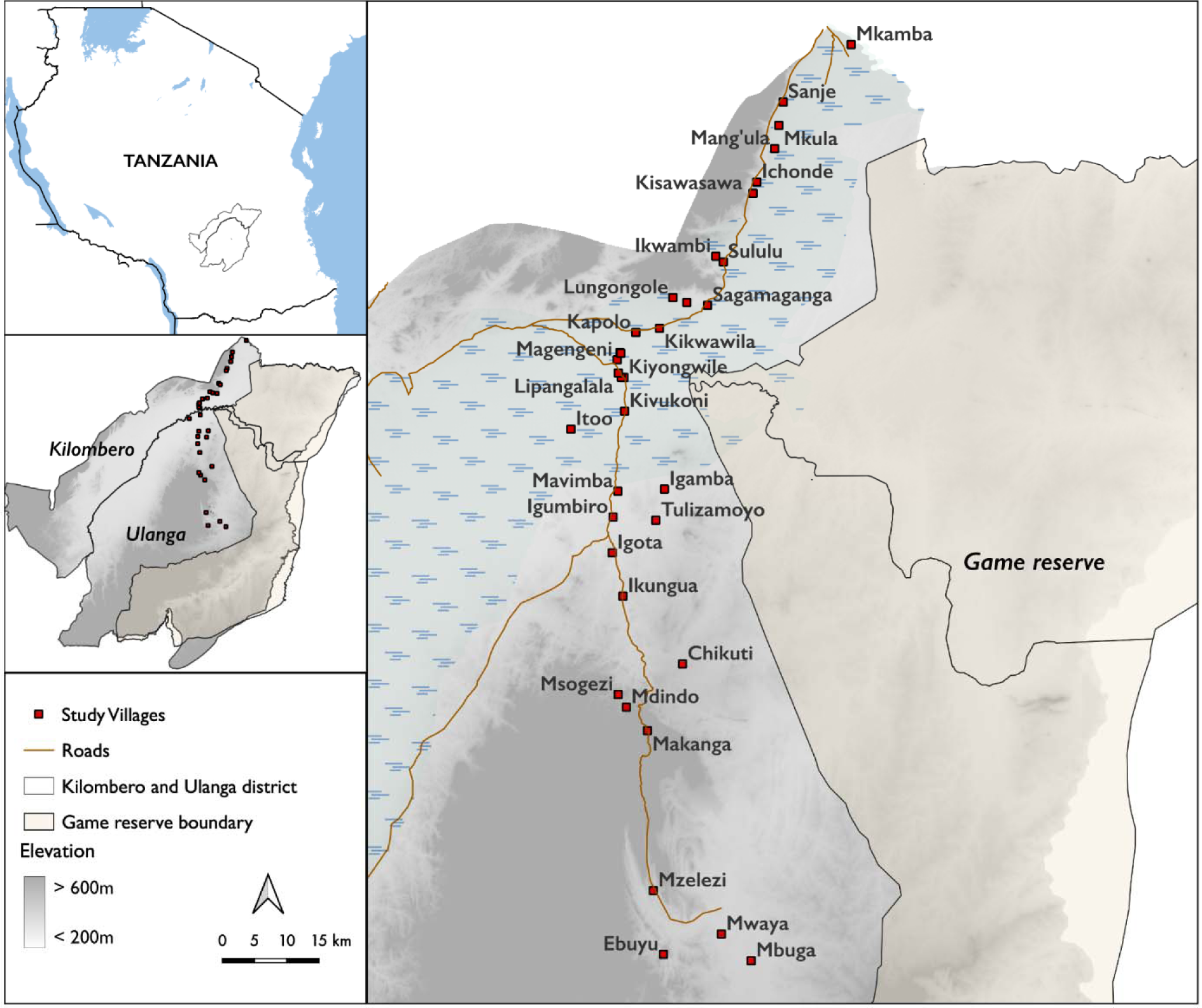
Study villages in Kilombero and Ulanga districts, south-eastern Tanzania.

### Study design, procedures and survey tools

The cross-sectional surveys were conducted over two consecutive years, from 2022 to 2023, covering the transition from the end of rainy seasons to the peak of dry seasons. Villages were randomly selected from each district, and sample sizes were proportionately determined based on the population of each village using the Cochran formula adjusted for finite populations [46–48]. The expected prevalence varied depending on the village and was derived from previous surveys and health centers within each village. Based on the estimated sample size mentioned above, representative households were selected by systematically randomizing the names of all households obtained from the respective village administrations.

The screening criteria included individuals aged 5-60 years who had not taken malaria medications in the preceding two weeks. This precaution aimed to prevent potential overestimation by RDTs, as they may detect residual traces post-treatment [49].

Individuals needing special medical attention, such as pregnant women, were excluded from the study. Each consenting individual undergoing malaria screening was assigned a unique identity number that was also linked to their corresponding household ID. On-site malaria diagnosis was conducted using: 1) RDTs via finger prick, 2) creating thick and thin blood smears, and 3) collecting 3-5 dried blood spots (cycles) on Whatman^TM^ filter paper cards. Subsequently, these samples were transported to the reference laboratory for microscopy and qPCR analysis. For children under five years, only screening with RDTs was done for prevalence estimations and not used for strata comparisons.

### Ethical considerations, survey team, and trainings

Permission to conduct this study was obtained from the Ifakara Health Institute Review Board (Ref: IHI/IRB/No: 1/2021) and the National Institute for Medical Research-NIMR (NIMR/HQ/R.8a/Vol. 1X/3735). Additionally, approvals were obtained from regional, district, ward, and respective selected village authorities before commencing the surveys, given the screening was done at centralized location in each village. Written informed consent was obtained from individual adult participants (and parents or guardians of those aged below 18) on the day before the actual testing. The study team consisted of 11 members, including three molecular laboratory technologists, four licensed medical laboratory microscopists, two licensed clinical officers, and two social scientists. Prior to the survey commencement, a five-day training session was conducted at the Ifakara Health Institute laboratory. This training covered explanations of the study protocols, pilot implementations, procedures for protecting human participants, quality assurance and training on data collection tools.

### Tests using malaria rapid diagnostic tests (RDTs)

A small blood drop obtained through a finger prick was collected onto the RDTs (SD Bioline Ag *Pf*/Pan), following the manufacturer’s instructions. The buffer solution was applied according to standard RDTs procedures and left on the bench surface for up to 20 minutes. The type RDT used were capable of detecting *P. falciparum* infections by targeting histidine-rich proteins 2, which react on the *Pf*-line. Additionally, they could detect *P. malariae* and *P. ovale* by targeting glycolytic lactate dehydrogenase, expressed by the Pan-line on RDTs [30]. The RDT results were recorded on a paper form, and any individuals who tested positive for malaria were promptly treated with Arthemether Lumefantrine (ALu), following Tanzania’s national malaria treatment guidelines [50].

### Tests using microscopy

Thick and thin blood smears were created in the field, stained with 10% Giemsa for 15 minutes then examined for the presence of malaria parasites under oil immersion at 100X magnification [47,51,52]. Two experienced microscopists independently read the slides, and discrepancies between them were resolved by a third, more experienced microscopist. They read the thick smear first, and if an infection was detected, the thin smear was read to identify parasite species. The presence of both asexual and sexual malaria parasite stages discriminating *P. falciparum*, *P. malariae*, and *P. ovale* was recorded. Asexual stage parasites were counted per 200 white blood cells and assuming 8000 WBC/μL [53]. The mean count of malaria parasite by microscopy between the two readers was calculated and confirmed by the third reader.

### Tests using real-time qPCR assays

A representative sample of approximately two thirds of all samples was randomly selected from each village and screened further by quantitative polymerase chain reaction (qPCR) i.e. 4905 samples out of the total 7628 samples. Out of five spots created on the Whatman^TM^ filter paper card, three spots of DBS were used for DNA extraction using Quick-DNA^TM^ Miniprep plus kit (Zymo Research,USA) [54], and eluted with50 μL of elution buffer, stored at −20°C for further detection and quantification of *P. falciparum* infections using probe-level allele-specific quantification (PlasQ)-multiplex qPCR assays protocols [37,55,56]. The detection and quantification of *P. falciparum* parasites were performed using the Bio-Rad CFX96 real-time PCR system (Bio-Rad Laboratories, USA) [55] and analyzed with Bio-Rad CFX maestro software. The qPCR reaction, PlasQ primers and probes mix, are summarized in supplementary online table 1& 2. DNA amplification processes included: activation at 95°C for 1 min, denaturation at 95°C for 15 seconds, and annealing and elongation at 57°C for 45 seconds for 45 cycles, followed by melting [55].

The qPCR assays were run with positive controls (samples with confirmed *P. falciparum*) and a non-template control (samples with no *P. falciparum* as negative control). For absolute parasite quantification, the WHO international standard for *P. falciparum* nucleic acid amplification techniques were used (WHO reference from NIBSC#04/176) [37]. The standard was reconstituted following the manufacturer’s instructions and serially diluted in the range of 100,000 parasites/μL to 0.01 and analyzed in triplicates.

During the qPCR assay, the prepared standards were run together with unknown samples, and at the end of the assay, the standard curve and samples were normalized and analyzed with Bio-Rad CFX maestro software. The obtained normalized Ct values of the samples and the linear regression equation derived from the standard curve were used to calculate the parasites density of the unknown samples, expressed as parasites per microlitre (parasites/μL) of blood.

### Data analysis

All results from RDTs, microscopy, and qPCR were entered into the Open Data Kit (ODK) system [57], and subsequently downloaded as an excel file for further cleaning. The datasets for RDTs, microscopy, and qPCR results were merged based on the participant’s ID using the Pandas Python package [58].

Generalized linear models (GLM) were used to evaluate the association between malaria infection risk and the variables of age and gender in a univariate analysis, run in Python using the stats model package [59]. Additionally, to evaluate the performance of RDTs and microscopy in fine-scale malaria stratifications compared to qPCR, their agreement was tested using Kappa statistic [60], and the resulting Kappa values interpreted as follows: κ < 0.20 as poor agreement, 0.21–0.40 as fair, 0.41–0.60 as moderate, 0.61–0.80 as substantially good and 0.81–1 as almost perfect agreement [61]. In addition, the positive predictive value (PPV) for RDTs and microscopy was computed, using qPCR results as the reference, per village, as (proportion of positive test results that are actually true positives, estimated as PPV = True Positives / (True Positives + False Positives)). Fine-scale stratification by villages using data from qPCR, RDT, and microscopy were used to generate the prevalence maps using quantitative Geographic Information System (qGIS) software, allowing visualization of malaria prevalence across villages. Additionally, the risk maps were created using inverse distance weighting (IDW) interpolation techniques. This method helped to estimate malaria risk by spatially interpolating the prevalence data, which provided a continuous surface of malaria risk across the study area.

The geometric mean of parasite density estimated by microscopy and qPCR was computed, and statistically compared the parasite densities between gender and age groups. The non-parametric Mann-Whitney statistics were used to compare the parasite densities between two categorical groups, while Kruskal-Wallis statistical tests were used to compare more than two categorical groups [62,63]. For example, differences in parasite densities between age groups were tested using Kruskal-Wallis statistics, and if statistically significant, the Mann-Whitney statistics were applied for pairwise statistical significance tests. All analyses that compared parasite densities excluded the negative class and focused solely on investigating parasite density distribution among malaria-positive patients. Lastly, to test for statistical correlations between parasite prevalence and parasite densities estimated by both qPCR and Microscopy, non-parametric Spearman’s rank correlation tests were employed [64]. Additionally, logistic regression model was also used to assess the probability of diagnostic tools to detect malaria infections at various parasite densities.

## Results

### Baseline study population

This survey covered 35 villages and 93 sub-villages across Ulanga and Kilombero districts. A total of 7,628 participants (>5 years) were recruited upon consent and tested for malaria using RDTs and microscopy, with 64.3% (4,905) of them also tested by qPCR (Figure 2). Males comprised 38% of the study population, while females made up 62%. Among the participants, 35% were school-aged children (5-15 years), and 65% were aged 16 years and above (Table 1). For children under five years, for whom screening was conducted with only RDTs to estimate the burden, the results are not reported here.

**Figure 2:**
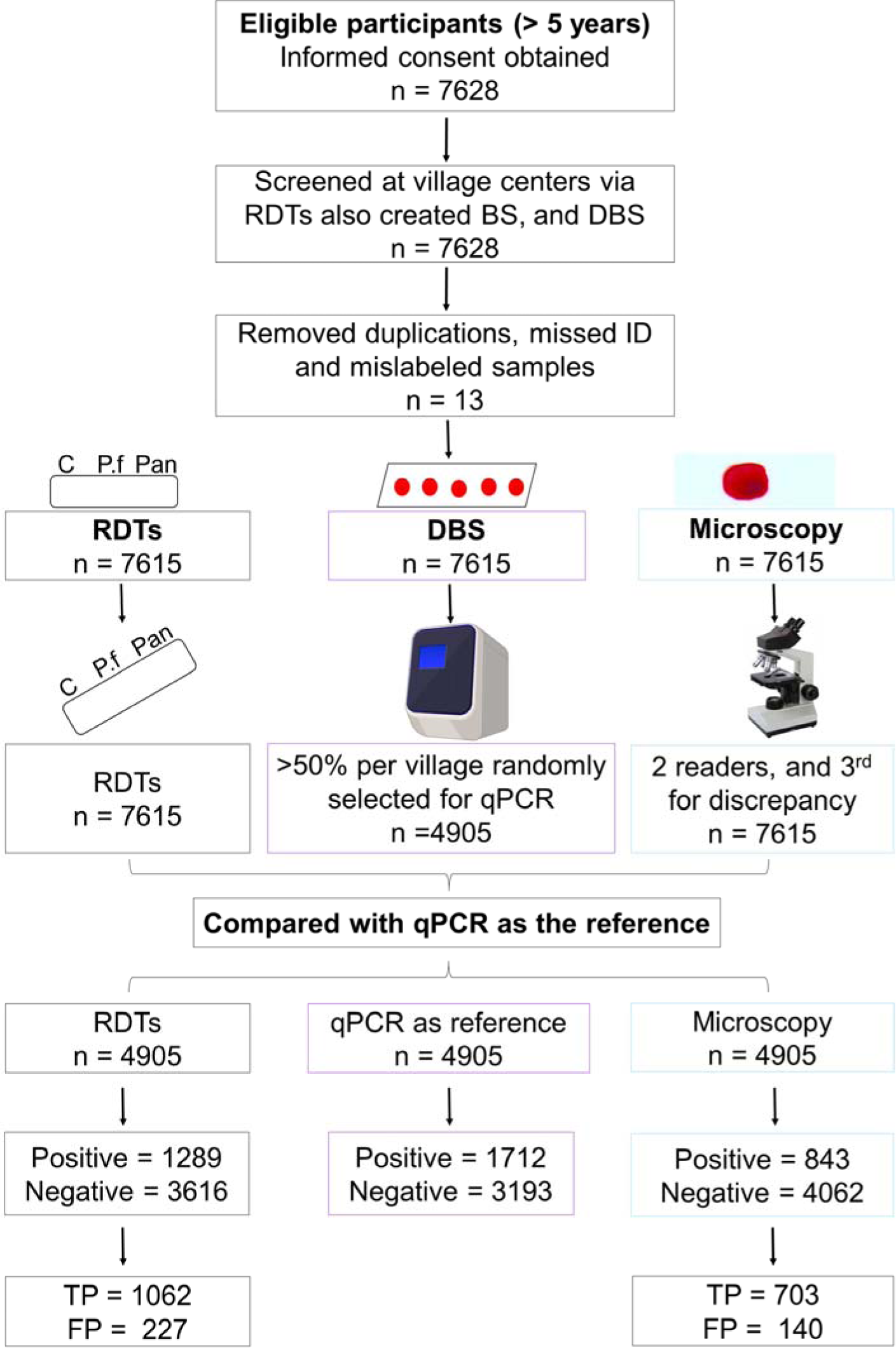
Schematic representation of the study sampling procedures

**Table 1:** Baseline characteristics of the study populations.

|  | **Kilombero District**<br>**n (%)** | **Ulanga District**<br>**n (%)** | **Total**<br>**N (%)** |
| --- | --- | --- | --- |
| **Villages** | 19 (54.3) | 16 (45.7) | 35.0 |
| **Sub Villages** | 48 (51.6) | 45 (48.4) | 93.0 |
| **Gender** |  |  |  |
| **Female** | 2164 (67.4) | 2573 (58.3) | 4737 (62.1) |
| **Male** | 1047 (32.6) | 1844 (41.7) | 2891 (37.9) |
| **Total** | **3211** | **4417** | **7628** |
| **Age Group** |  |  |  |
| **5-10 years** | 519 (16.2) | 935 (21.2) | 1454 (19.1) |
| **11-15 years** | 400 (12.5) | 802 (18.2) | 1202 (15.8) |
| **16-20 years** | 198 (6.2) | 333 (7.5) | 531 (7.0) |
| **>20 years** | 2094 (65.2) | 2347 (53.1) | 4441 (58.2) |
| **Total** | **3211** | **4417** | **7628** |

### Malaria prevalence by RDTs, Microscopy and qPCR

In the Ulanga district, malaria transmission was found to be high by both qPCR and RDTs, with *P. falciparum* prevalence rates of 53.89% [95% CI, 52.06-55.72] and 38.35% [95% CI, 36.92-39.79] respectively. However, microscopy categorized it as moderate, with a prevalence rate of 26.07% [95% CI: 24.77-27.36] (Table 2). Within this moderate to high transmission strata in Ulanga, males had a significantly higher prevalence of malaria compared to females. The odds of malaria infection in males compared to females were estimated as 1.54 [95% CI: 1.36-1.74] (P < 0.001) by RDTs, 1.45 [95% CI: 1.27-1.66] (P < 0.001) by microscopy, and 1.54 [95% CI: 1.36-1.74] (P < 0.001) by qPCR (Table 2). All tests - RDTs, microscopy, and qPCR - indicated that school-age children (5-15 years) had a significantly higher prevalence of malaria infections than the other age groups (Table 2).

**Table 2:** Malaria prevalence in Ulanga district by sex and age groups, as measured using RDTs, microscopy and qPCR.

| Attribute | RDTs |  |  |  | Microscopy |  |  |  | qPCR |  |  |  |
| --- | --- | --- | --- | --- | --- | --- | --- | --- | --- | --- | --- | --- |
|  | Positive/<br>Total<br>tested | Prevalence<br>(%)<br>[95% CI] | Odds of<br>infection<br>[95% CI] | P-value | Positive/<br>Total<br>tested | Prevalence<br>(%)<br>[95% CI] | Odds of<br>infection<br>[95% CI] | P-value | Positive/<br>Total<br>tested | Prevalence<br>(%)<br>[95% CI] | Odds of<br>infection<br>[95% CI] | P-value |
| **Overall prevalence** | 1689/4404 | 38.4<br>[36.9-39.8] | - | - | 1148/4404 | 26.1<br>[24.8 – 27.4] | - | - | 1531/2841 | 53.9<br>[52.1-55.7] | - | - |
| **Sex** |  |  |  |  |  |  |  |  |  |  |  |  |
| **Female** | 874/2566 | 34.1<br>[32.2-35.9] | Ref |  | 593/2566 | 23.1<br>[21.5 – 24.7] | Ref | - | 830/1659 | 50.0<br>[47.6-52.4] | Ref | - |
| **Male** | 815/1838 | 44.3<br>[42.1-46.6] | 1.5<br>[1.4-1.7] | <0.001 | 555/1838 | 30.2<br>[28.1-32.3] | 1.4<br>[1.3-1.6] | <0.001 | 701/1182 | 59.3<br>[56.5-62.1] | 1.5<br>[1.4-1.8] | <0.001 |
| **Age group** |  |  |  |  |  |  |  |  |  |  |  |  |
| **5-10 years** | 497/933 | 53.3<br>[50.1-56.5] | Ref |  | 341/933 | 36.6<br>[33.5-39.6] | Ref | - | 338/622 | 54.3<br>[50.4-58.3] | Ref | - |
| **11-15 years** | 479/801 | 59.8<br>[56.4-63.2] | 1.3<br>[1.1-1.6] | 0.006 | 358/801 | 44.7<br>[41.3-48.1] | 1.4<br>[1.2-1.7] | <0.001 | 355/534 | 66.5<br>[62.5-70.5] | 1.3<br>[1.1-1.6] | 0.006 |
| **16-20 years** | 154/332 | 46.4<br>[41.0-51.8] | 0.8<br>[0.6-1.0] | 0.031 | 109/332 | 32.8<br>[27.9-37.9] | 0.9<br>[0.7-1.1] | 0.224 | 127/210 | 60.5<br>[53.9-67.1] | 0.8<br>[0.6-1.0] | 0.031 |
| **>20 years** | 559/2338 | 23.9<br>[22.2-25.6] | 0.3<br>[0.2-0.4] | <0.001 | 340/2338 | 14.5<br>[13.1-16.0] | 0.3<br>[0.3-0.4] | <0.001 | 711/1475 | 48.2<br>[45.7-50.8] | 0.3<br>[0.2-0.3] | <0.001 |

In Ifakara council, within the Kilombero district, both RDTs and microscopy categorized the area as a low risk stratum, with observed prevalence rates of 2.68 [95% CI: 2.12-3.24] and 1.84 [95% CI: 1.37 – 2.30], respectively (Table 3). However, qPCR classified Kilombero district as a moderate risk stratum with a prevalence rate of 8.77 [95% CI: 7.55-9.99] (Table 3). Notably, there were no statistically significant differences in malaria infection risk between males and females in this low to moderate transmission setting, as indicated by both RDTs (Odds prevalence 1.51% [95% CI; 0.97, 2.33], P=0.065) and microscopy (Odds prevalence 1.33% [95% CI; 0.78, 2.25], P=0.293), as well as qPCR (Odds prevalence 1.09% [95% CI: [0.80-5.67], P = 0.567). Additionally, school age children (5-15 years) exhibited a significantly higher risk of malaria infections compared to those above 15 years old, as demonstrated by both RDTs, microscopy, and PCR (Table 3).

**Table 3:** Malaria prevalence in the Kilombero district by sex and age groups, as measured using RDTs, microscopy and qPCR.

| Attribute | RDTs |  |  |  | Microscopy |  |  |  | qPCR |  |  |  |
| --- | --- | --- | --- | --- | --- | --- | --- | --- | --- | --- | --- | --- |
|  | Positive/<br>Total<br>tested | Prevalence<br>(%)<br>[95% CI] | Odds of<br>infection<br>[95% CI] | P-value | Positive/<br>Total<br>tested | Prevalence<br>(%)<br>[95% CI] | Odds of<br>infection<br>[95% CI] | P-value | Positive/<br>Total<br>tested | Prevalence<br>(%)<br>[95% CI] | Odds of<br>infection<br>[95% CI] | P-value |
| **Overall prevalence** | 86/3211 | 2.7<br>[2.1-3.2] | - | - | 59/3211 | 1.8<br>[1.4 – 2.3] | - | - | 181/2064 | 8.8<br>[7.6-10.0] | - | - |
| **Sex** |  |  |  |  |  |  |  |  |  |  |  |  |
| **Female** | 50/2164 | 2.3<br>[1.7-2.9] | Ref | - | 36/2164 | 1.2<br>[1.1-2.2] | Ref |  | 118/1385 | 8.5<br>[7.1-10.0] | Ref | - |
| **Male** | 36/1047 | 3.4<br>[2.3-4.5] | 1.5<br>[0.9-2.3] | 0.065 | 23/1047 | 2.2<br>[1.3 – 3.1] | 1.3<br>[0.8-2.3] | 0.293 | 63/679 | 9.3<br>[7.1-11.5] | 1.1<br>[0.8-1.5] | 0.567 |
| **Age group** |  |  |  |  |  |  |  |  |  |  |  |  |
| **5-10 years** | 21/519 | 4.1 [2.4-5.7] | Ref | - | 16/519 | 3.1<br>[1.6-4.6] | Ref | - | 33/343 | 9.6<br>[6.5-12.7] | Ref | - |
| **11-15 years** | 17/400 | 4.3<br>[2.3-6.2] | 1.1<br>[0.6-2.0] | 0.877 | 10/400 | 2.50<br>[1.0-4.03] | 0.8<br>[0.4-1.8] | 0.597 | 28/256 | 10.9<br>[7.1-14.8] | 1.2<br>[0.7-2.0] | 0.59 |
| **16-20 years** | 9/198 | 4.6<br>[1.6-7.5] | 1.1<br>[0.5-2.5] | 0.765 | 4/198 | 2.02<br>[0.1-3.4] | 0.6<br>[0.2-2.0] | 0.443 | 16/123 | 13.0<br>[7.1-19.0] | 1.4<br>[0.7-2.7] | 0.295 |
| **>20 years** | 39/2094 | 1.9<br>[1.3-2.4] | 0.5<br>[0.3-0.8] | 0.003 | 29/2094 | 1.38<br>[0.88-1.89] | 0.4<br>[0.2-0.8] | 0.009 | 104/1342 | 7.8<br>[6.3-9.2] | 0.8<br>[0.5-1.2] | 0.259 |

### Micro-stratification of malaria risk using data collected by qPCR, RDTs, and microscopy

Significant variability in malaria infections was observed at the individual village level, with prevalence rates ranging from 0% to over 50% across the study area (Figure 3 & Table 4). Additionally, the method used to test for malaria significantly impacted the risk categorization of villages. Among the 35 villages surveyed, qPCR data indicated that only one village (1.2% of all villages) had very low malaria prevalence (PfPR < 1%). In contrast, RDTs identified 12 villages (34.3% of all villages) and microscopy identified 11 villages (31.4% of all villages) as having very low prevalence. For moderate transmission, qPCR, RDTs, and microscopy categorized 15, 9, and 8 villages, respectively. For high transmission, qPCR identified 14 villages, RDTs identified 8, and microscopy identified 6. Notably, qPCR detected more malaria infections than RDTs and microscopy, resulting in many villages being classified into higher transmission categories.

**Figure 3:**
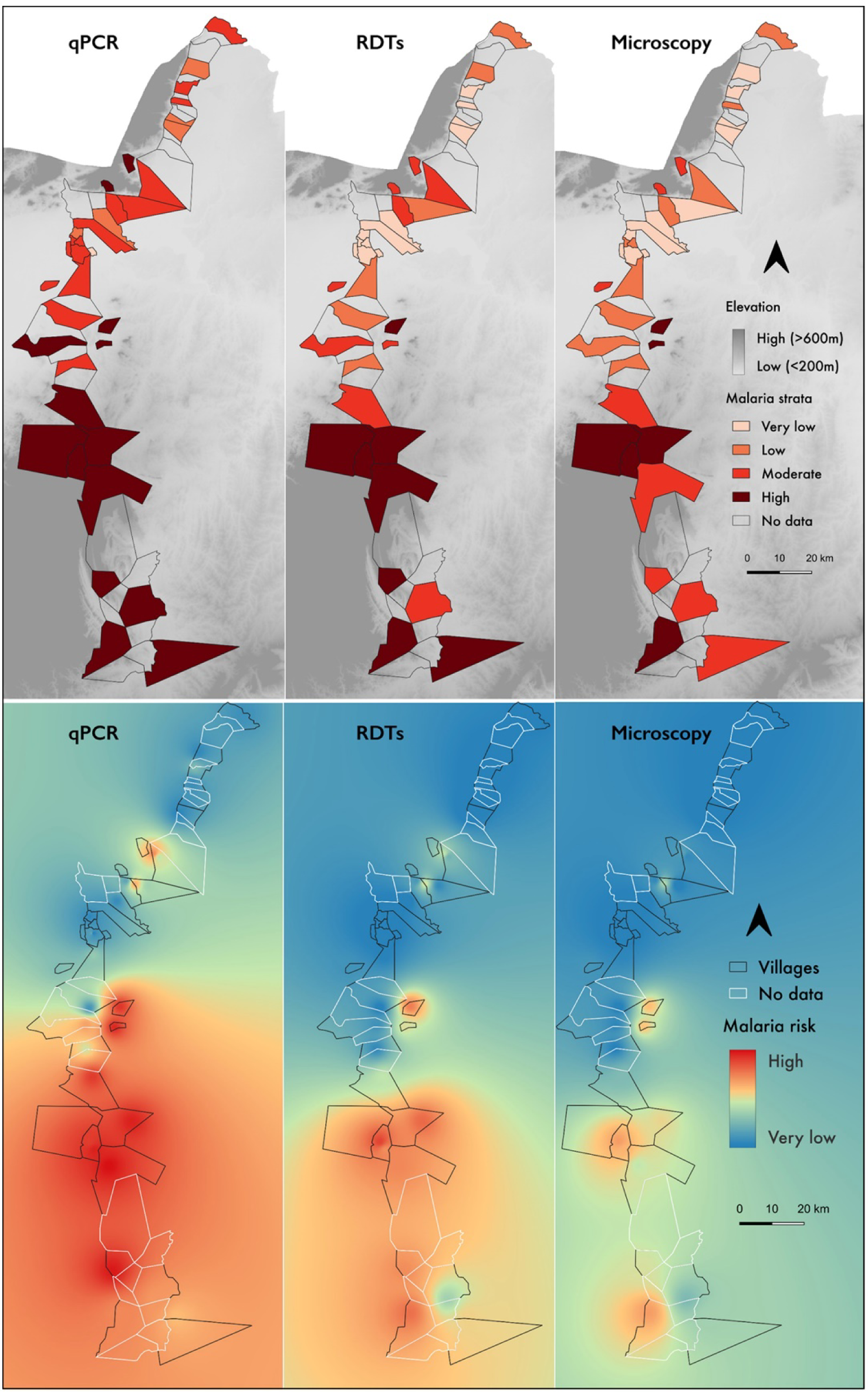
Fine-scale malaria mapping of 35 surveyed villages in the Ulanga and Kilombero districts using qPCR, RDTs, and microscopy data is shown in the top panel. The bottom panel indicates malaria risk generated by interpolating prevalence data obtained for each surveyed village by qPCR, RDTs, and Microscopy.

**Table 4:** Number of villages categorized into different risk strata based on the P. falciparum prevalence rate (PfPR) data from qPCR, RDTs and microscopy.

| Risk strata | Prevalence | Number of villages |  |  |  |  |  |
| --- | --- | --- | --- | --- | --- | --- | --- |
|  |  | qPCR |  | RDTs |  | Microscopy |  |
|  |  | Count | % | No. | % | No. | % |
| Very low | PfPR < 1% | 1 | 2.9 | 12 | 34.3 | 11 | 31.4 |
| Low | 1% ≤ PfPR < 5% | 5 | 14.3 | 6 | 17.1 | 10 | 28.6 |
| Moderate | 5% ≤ PfPR < 30% | 15 | 42.9 | 9 | 25.7 | 8 | 22.9 |
| High | PfPR ≥ 30% | 14 | 40.0 | 8 | 22.9 | 6 | 17.1 |
| Total |  | 35 | 100.0 | 35 | 100.0 | 35 | 100.0 |

Overall, using qPCR data, over 80% of the villages were classified as moderate to high risk, significantly higher than the 48% classified by RDTs and 40% by microscopy. Conversely, while only 17% of the villages were classifiable as having low or very low malaria risk based on qPCR data, as high as 51% and 60% of the villages were classified into these same categories based on RDT and microscopy data (Table 4 and Figure 4)

**Figure 4:**
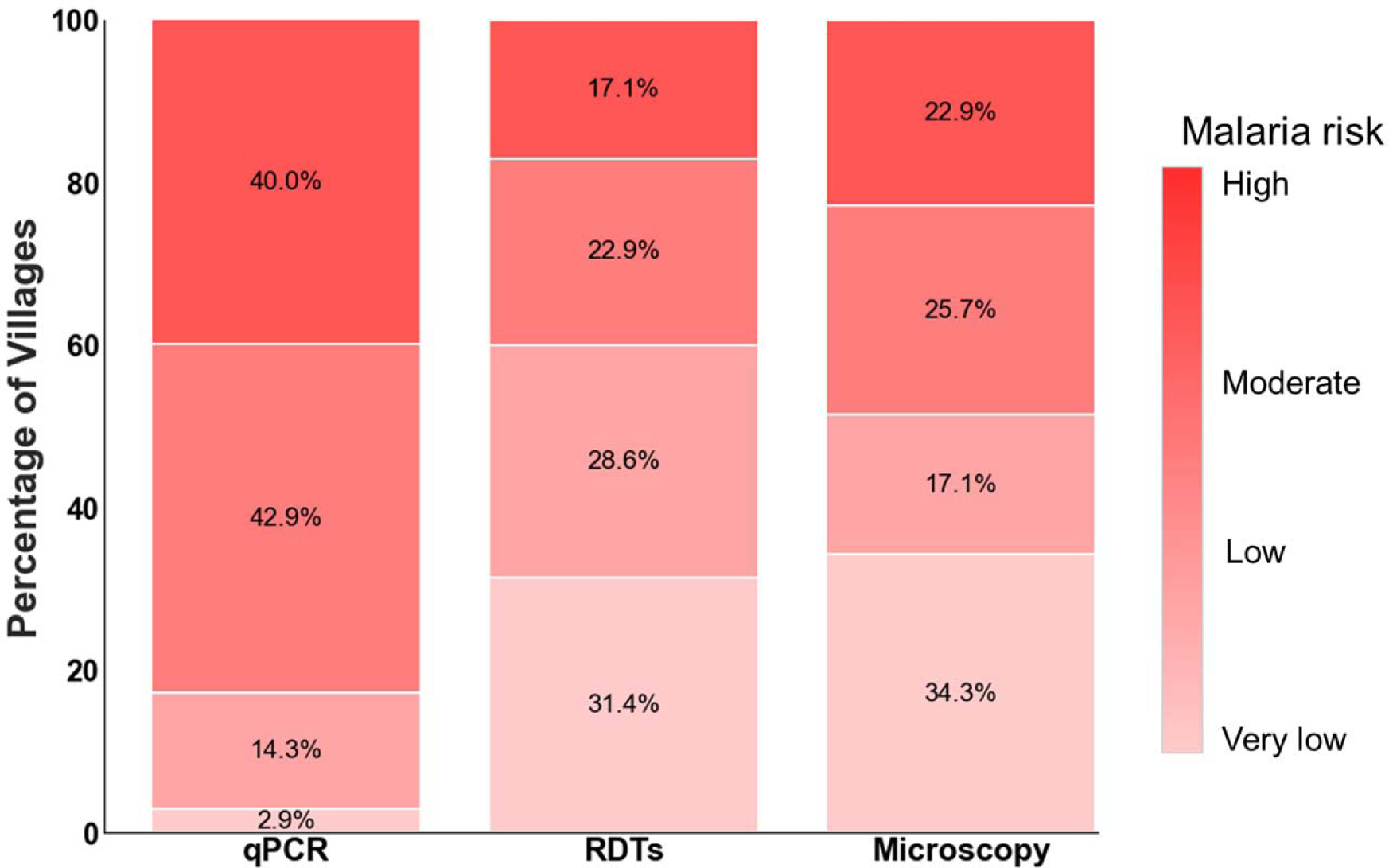
Percentage of villages categorized by different testing methods as either very low risk, low risk, moderate risk or high risk (total number of villages = 35).

### Comparison of the perfomance of RDTs and microscopy relative to qPCR

In this comparative analysis, only samples tested by all three methods—PCR, RDTs, and microscopy—were included, totaling 64.3% (n = 4905) of the samples (Table 5). Among these, qPCR identified 1712 (34.9%) as positive, whereas RDTs and microscopy classified 1289 (26.3%) and 843 (17.2%) positives, respectively (Table 5). A substantial number of positive samples, otherwise detectable by qPCR, were missed by both RDTs and microscopy, suggesting the existence of subpatent infections (table 5 and Figure 5). Of all samples analyzed by qPCR, RDTs, and microscopy, 647 out of 4,905 (13.2%) were categorized as positive by all three methods (Figure 5).

**Figure 5:**
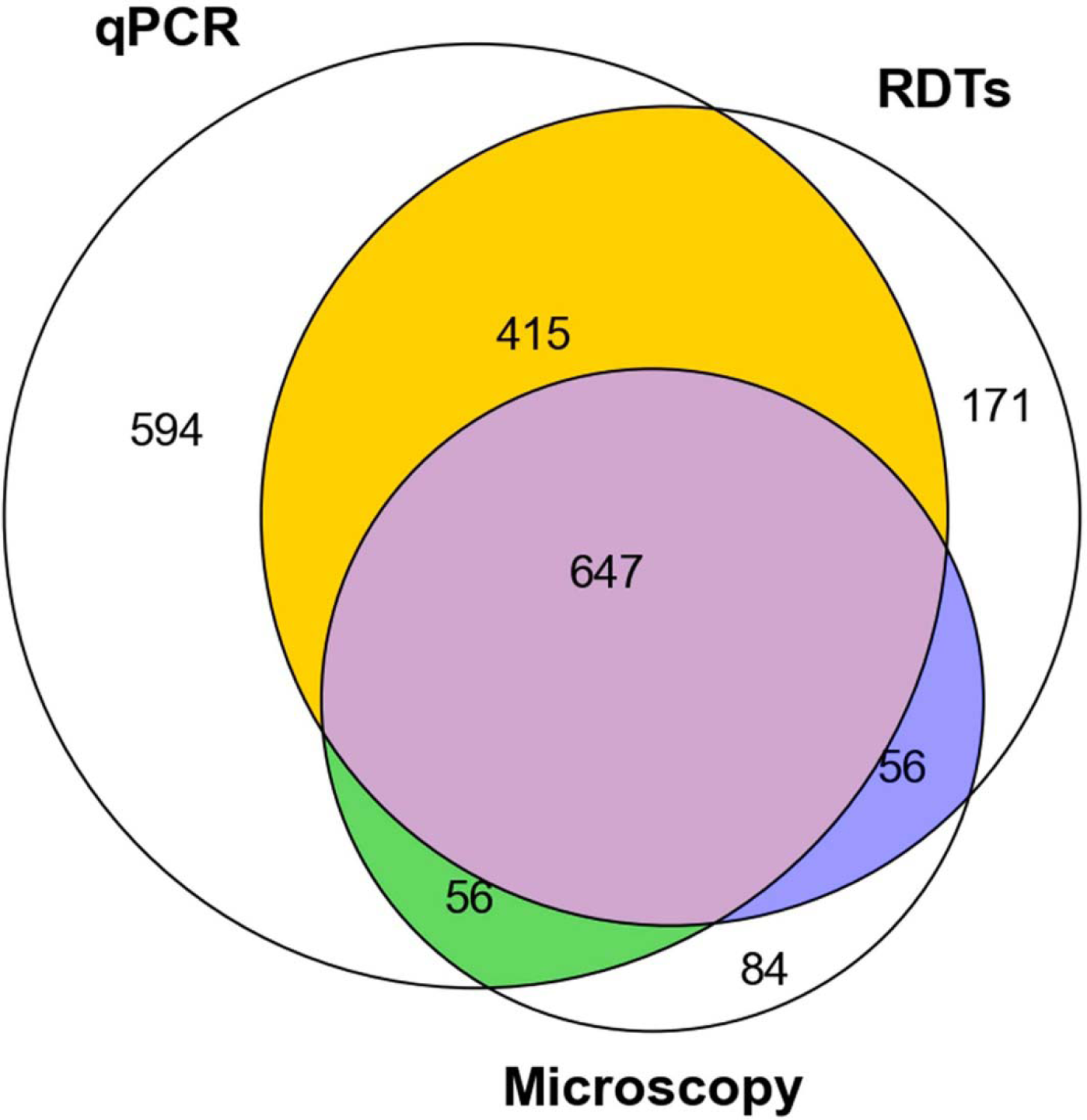
The Venn diagram illustrates positive samples detected exclusively by a specific tool while the other two missed them (qPCR only: 594 positive, RDT only: 171 positive, Microscopy only: 84 positive). Additionally, it shows intersections indicating positive detection by two tools when one detects negative (qPCR & RDT: 415 positive; qPCR & Microscopy: 56 positive; RDT & Microscopy: 56 positive). It also indicates intersections where all tools detect positive samples (qPCR, RDT, & Microscopy: 647 positive).

**Table 5:**
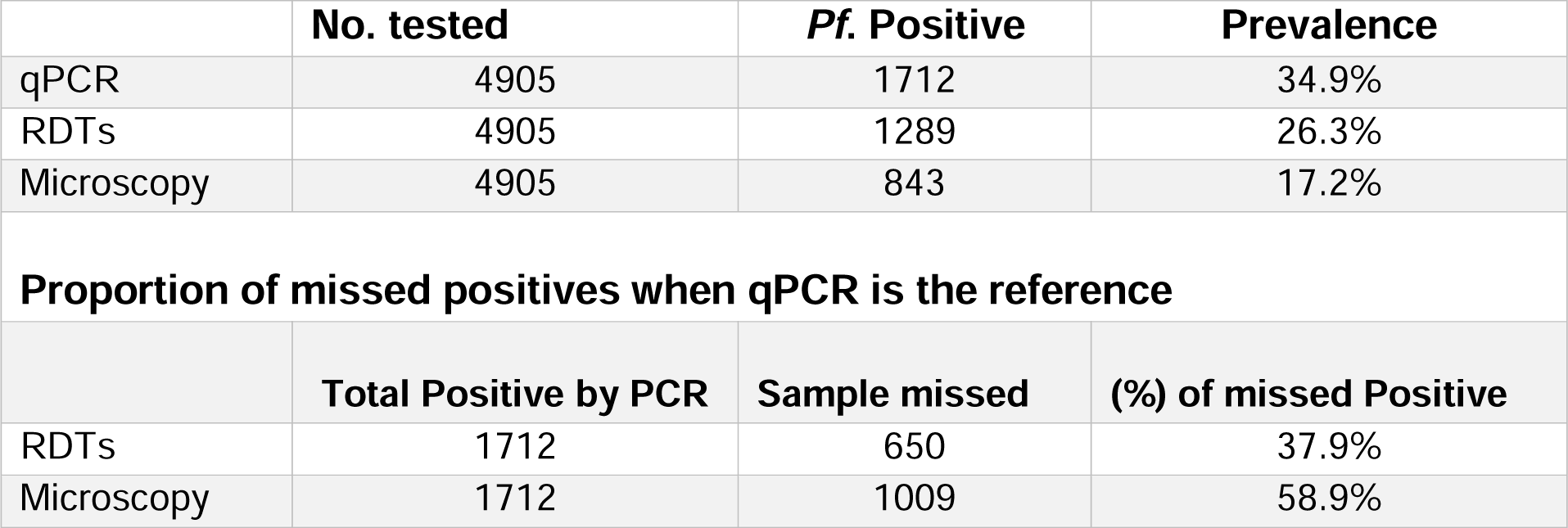
Overall prevalence in the 35 surveyed villages estimated using qPCR, RDTs, and microscopy. Additionally, the table summarizes the proportion of malaria positive samples missed by RDTs and microscopy when qPCR is used as the reference.

|  | **No. tested** | ***Pf.* Positive** | **Prevalence** |
| --- | --- | --- | --- |
| qPCR | 4905 | 1712 | 34.9% |
| RDTs | 4905 | 1289 | 26.3% |
| Microscopy | 4905 | 843 | 17.2% |
| **Proportion of missed positives when qPCR is the reference** |  |  |  |
|  | **Total Positive by PCR** | **Sample missed** | **(%) of missed Positive** |
| RDTs | 1712 | 650 | 37.9% |
| Microscopy | 1712 | 1009 | 58.9% |

Interestingly, qPCR classified 56 out of 4,905 (1.1%) samples as negative, while both RDTs and microscopy agreed they were positive (Figure 5). Similarly, the same proportion was agreed upon by qPCR and microscopy as positive, while RDTs classified them as negative. Microscopy classified 415 out of 4,905 (8.5%) samples as negative, while both RDTs and qPCR agreed they were positive.

Both RDT and microscopy missed several infections otherwise identified by qPCR. This category of false negatives included cases where qPCR identified a sample as positive, but microscopy identified it as negative, cases classified as positive by qPCR but negative by RDTs, and cases where RDTs indicated positive results while microscopy indicated negative result. Out of the 1712 positives detected by qPCR, RDTs missed 650 (37.97%) and microscopy missed 1009 (58.9%) (Table 5). Additionally, when comparing microscopy to RDTs, microscopy failed to detect 45.46% (586/1289) of malaria infections detected by RDTs. RDTs correctly identified 1062 (62.03%) samples as true positives, while microscopy identified 703 (41.06%) as true positives (Table 6). Furthermore, RDTs misclassified 227 (7.10%) samples as false positives, while microscopy misclassified 140 (4.38%) (Table 6).

**Table 6:** Evaluation metrics for assessing the performance of RDTs and microscopy relative to qPCR during the fine-scale stratification of malaria risk in Ulanga and Kilombero districts, south-eastern Tanzania.

| Test characteristics | RDTs |  | Microscopy (Thick smear) |  |
| --- | --- | --- | --- | --- |
|  | Evidence from Published studies | Evidence from this study | Evidence from Published studies | Evidence from this study |
| **True Positives (PCR positive=1712)** | - | 1062 | - | 703 |
| **False Positives (PCR negative)** | - | 227 | - | 140 |
| **True Negatives (PCR negative=3193)** | - | 2966 | - | 3057 |
| **False Negatives (PCR positive)** | - | 650 | - | 1009 |
| **Sensitivity [95% CI]** | 56 - 78%<br>[55,65–68] | 62.0%<br>[95 CI: 60.0-64.2] | 31-63%<br>[55,65–68] | 41.0%<br>[95 CI: 38.8-43.4] |
| **Specificity [95% CI]** | 91-99%<br>[55,65–68] | 92.9%<br>[95 CI: 92.0-93.7] | 82-100%<br>[55,65–68] | 95.6%<br>[95 CI: 94.9-96.3] |
| **Positive Predictive Value [95% CI]** | 75 -100<br>[69,70] | 82.4%<br>[95 CI: 80.3 -84.4] | 70 -100<br>[66,69] | 83.4%<br>[95 CI: 80.8-86.0] |
| **Negative Predictive Value [95% CI]** | 75-99<br>[66,68,71] | 82.0%<br>[95 CI: 80.7-83.2] | 66-99<br>[66,68] | 75.2%<br>[95 CI: 73.8-76.4] |
| **Kappa value [95% CI]** | 50-77<br>[55,65–68] | 0.6<br>[95 CI: 0.6-0.6] | 54% [48-60]<br>[55,65–68] | 0.4<br>[95 CI: 0.4-0.40] |
| **Accuracy** | - | 82.1% | - | 76.57% |

### Positive predictive values (PPVs), sensitivity, specificity, and agreement of RDTs and microscopy when compared to qPCR

Considering qPCR as the benchmark, the sensitivity (the proportion of actual positives which were correctly identified as such) of RDTs was 62.0% [95% CI: 60.0-64.2], while that of microscopy was 41.0% [95% CI: 38.8 - 43.4]. The specificity (proportion of actual negatives which were correctly identified as such) was 92.9% [95% CI: 92.00-93.7] for RDTs and 95.6% [95% CI: 94.9-96.3] for microscopy (Table 6). Overall, the positive predictive value (PPV), i.e. the probability that individuals with a positive test result actually have the disease, was 82.4% [95% CI: 80.3-84.4] for RDTs and 83.4% [95% CI: 80.8-86.0] for microscopy (Table 6). Importantly however, the PPV for both RDTs and microscopy varied with malaria endemicity, generally increasing with prevalence, ranging from less than 20% in very low transmission areas to over 80% in high transmission areas (Figure 06).

When considering the micro-strata generated using qPCR data, the PPV of RDTs and microscopy started at 0% in very low risk strata and gradually increased to >80% as villages shifted towards high risk strata (Figure 6A). However, when referring to the strata generated using RDTs data (Figure 6B), the PPV of both RDTs and microscopy started at 20% in very low risk strata and gradually increased to >80% in high-risk strata. The agreement between RDTs and qPCR was good (Kappa value = 0.58 [95% CI: 0.56-0.61]), while the agreement between microscopy and qPCR showed fair agreement (Kappa value = 0.42 [95%CI: 0.39-0.44]), (Table 6).

**Figure 6:**
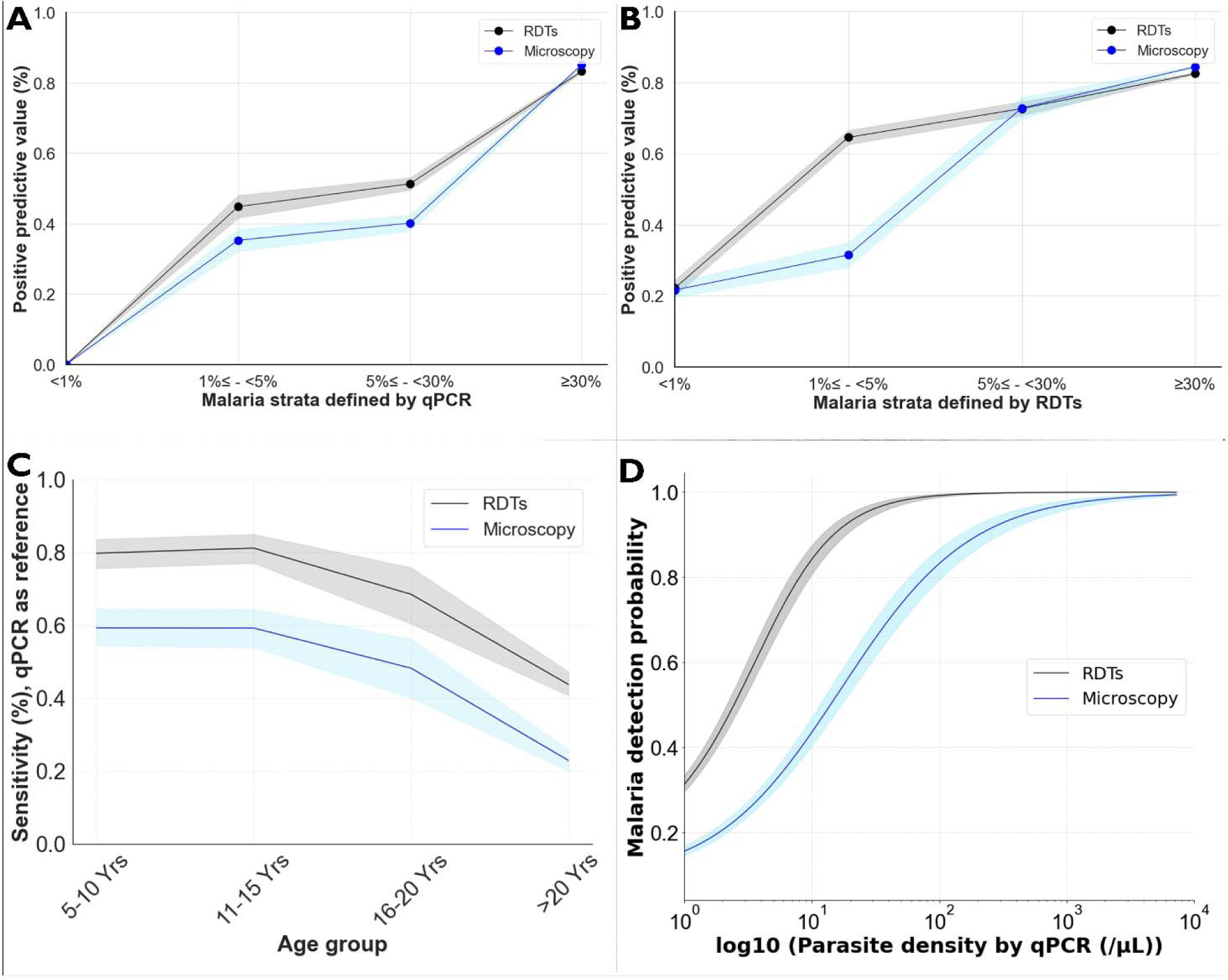
Estimates of the positive predictive values (PPVs) of RDTs and microscopy at different malaria endemicities across the study villages, defined based on either qPCR data (A) or RDT data (B). Panel C) illustrates the trend of sensitivity of both RDTs and microscopy relative to age groups. Panel D) displays the detection probability of both RDTs and microscopy relative to the parasite density estimated by qPCR.

The sensitivity of both RDTs and microscopy varied by age, where RDTs sensitivity was higher for school-aged children (>80%) and dropped to 75% and 60% for 16-20 years and >20 years, respectively (Figure 6C). A similar trend of sensitivity was observed for microscopy (Figure 6C), indicating that RDTs and microscopy perform better in detecting malaria in school-aged children compared to adults.

The relationship between parasite density and the malaria detection probability by RDTs and microscopy was also examined. In this analysis, the probability of RDTs detecting positive malaria infections was maximized reaching 1 at 100 parasites/μL, where the logistic regression (logit (p)) model saturated (Figure 6D). At this density in contrast, the probability of microscopy to detect malaria infections was only 0.85% (Figure 6D), suggesting higher sensitivity of RDTs vs microscopy.

### Parasites density estimates and their correlations with *Plasmodium* prevalence

Further, asexual parasite densities estimated by both microscopy and qPCR were investigated and compared across different sex and age groups using Mann-Whitney statistics for two categories and the Kruskal-Wallis statistical test for more than two categories. Overall, PCR was capable of detecting approximately 100 fold lower parasite densities compared to microscopy. The geometric mean asexual parasite density estimated by microscopy was 2206.4 parasites/μL (95% CI: 1976.7-2462.8), while that estimated by PCR was 27.07 parasites/μL (95% CI: 23.23-31.54) (Table 7).

**Table 7:** Parasite densities by gender and age as estimated using Microscopy slides and quantitative PCR assays.

|  | Microscopy |  |  | qPCR |  |  |
| --- | --- | --- | --- | --- | --- | --- |
|  | Geometric mean (/μL) | 95% CI (/μL) | P-value | Geometric mean (/μL) | 95% CI (/μL) | P-value |
| **Overall parasite density** | 2206.39 | 1976.68-2462.81 | - | 27.07 | 23.23-31.54 | - |
| **Sex** |  |  |  |  |  |  |
| **Female** | 2085.06 | 1775.94-2447.98 | **0.11<sup>a</sup>** | 20.48 | 16.76-25.02 | **<0.001<sup>a</sup>** |
| **Male** | 2341.22 | 2014.74-2720.61 |  | 38.32 | 30.33-48.41 |  |
| **Age group** |  |  |  |  |  |  |
| **5-10 years** | 3715.39 | 3040.63-4534.91 | **<0.001<sup>b</sup>** | 108.32 | 74.55-157.42 | **<0.001<sup>b</sup>** |
| **11-15 years** | 2398.92 | 2017.28—<br>2852.75 |  | 76.83 | 55.26-106.83 |  |
| **16-20 years** | 2117.02 | 1443.62-3104.53 |  | 30.22 | 17.47-52.25 |  |
| **>20 years** | 1182 | 968.15-1444.34 |  | 9.05 | 7.59-10.79 |  |

**a=** Mann-Whitney Test
**b=** Kruskal-Wallis Test

The asexual parasite density of infected individuals significantly differed between males and females as estimated by qPCR (P < 0.001), with males harboring a higher parasite density compared to females, though similar trend was observed by microscopy, this sex difference was not statistically detectable by microscopy (P = 0.11). Importantly, the geometric mean parasite density estimated by both microscopy and qPCR per village demonstrated a significant positive correlation with the parasite prevalence of the respective village. Thus, villages with high malaria prevalence also had high malaria parasite densities compared to villages with lower prevalence (Figure 7C-F). Considering qPCR-estimated geometric mean parasite densities, the Spearman rank correlation score was 0.77 (P < 0.001) and 0.76 (P < 0.001) when the malaria prevalence of the villages was estimated by qPCR and RDTs, respectively (Figure 7C & 7E). On the other hand, the Spearman rank correlation for the parasite density estimated by microscopy was 0.55 (P < 0.003) and 0.48 (P < 0.012) for qPCR and RDTs estimated prevalence of the villages, respectively (Figure 7D & 7F).

**Figure 7:**
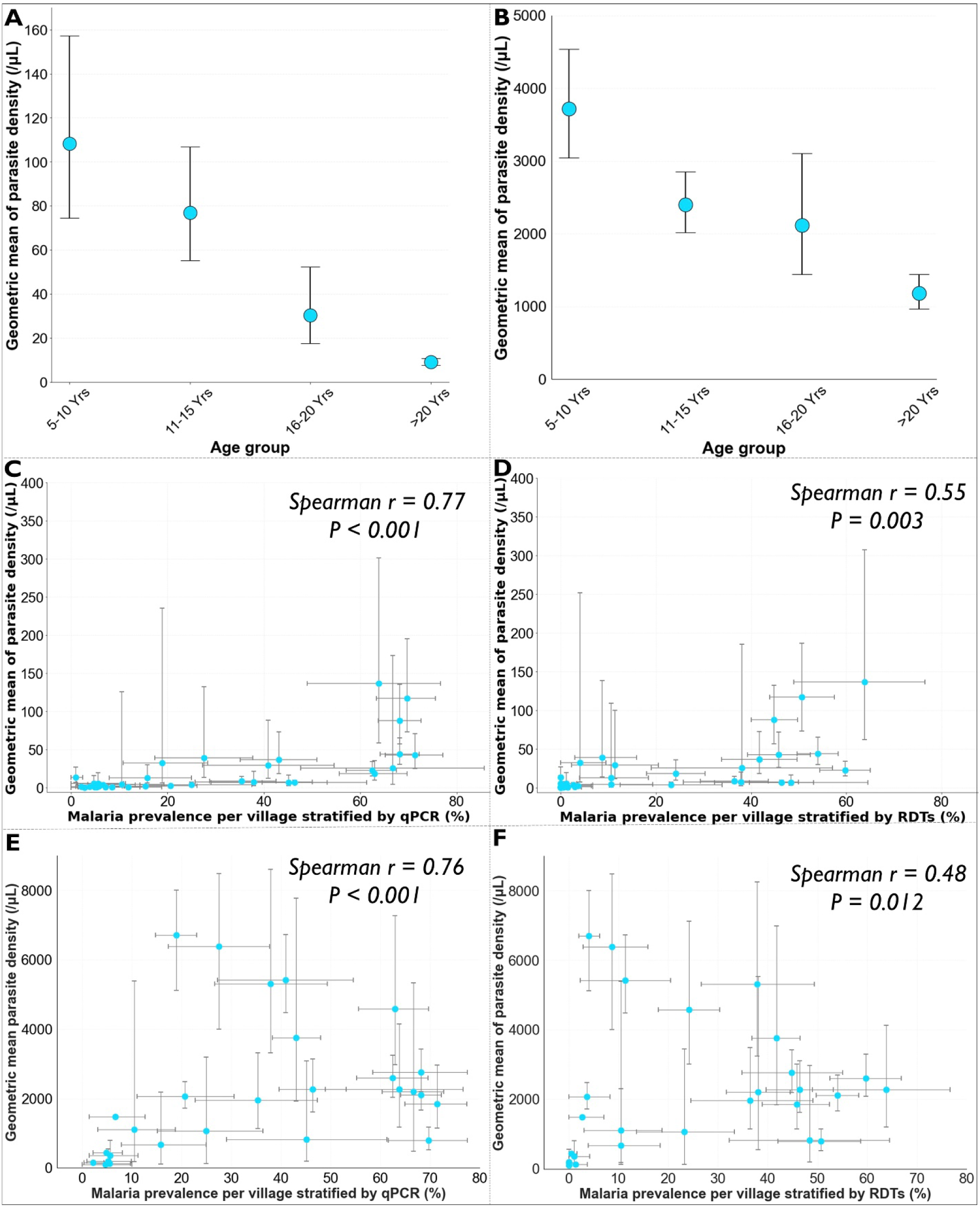
Geometric mean parasite densities per age group estimated by A) qPCR and B) microscopy, and correlations with parasite prevalence (C-F).

When parasite density by age groups were analyzed, both microscopy and qPCR revealed a significant difference in estimated malaria parasite densities between age groups based on Kruskal-Wallis statistics (P < 0.001) (Table 7). Pairwise tests by Mann-Whitney statistics revealed that school-aged children (5-15 years old) harbored a higher parasite density than those 16 years old and above (P < 0.001) as indicated by both microscopy and qPCR (Figure 7A & 7B).

## Discussion

In malaria-endemic countries, data-driven risk stratification is increasingly used at district or higher regional levels to guide intervention strategies and optimize resource allocation. Additionally, the geographical variations in levels of endemicities and the shift towards elimination in some settings necessitates finer resolution for optimal resource allocation [6,10,72]. In most settings in Africa, the data used for epidemiological stratification of malaria typically comes from rapid diagnostic tests (RDTs) or microscopy-based testing, which despite wide scale availability and low operational costs, often perform poorly in low transmission settings [41,73,74]. While direct comparisons of these diagnostic tools for fine-scale stratification are currently limited, selecting the most appropriate data sources and testing methods is crucial, as different methods can yield significantly different results depending on endemicity, particularly in elimination settings. Even without alternative testing methods, data users and decision-makers need to understand the limitations of their selected approaches, especially the weaknesses of current dominant data sources like RDTs or microscopy. In this study, we conducted a high-resolution survey of *P. falciparum* malaria in two Tanzanian districts, comparing fine-scale strata obtained using RDTs, microscopy, and qPCR assays. The findings will provide the necessary evidence to the malaria programs at national and district levels on how RDTs, microscopy, and qPCR compare when used at local levels. Moreover, the findings demonstrate that selection of appropriate test methods is essential for more precise resource allocation and improved understanding of disease patterns.

The findings highlight significant variability in malaria risk at a fine scale. Within less than 150 kilometers, malaria prevalence estimates ranged from 0% to over 50% across contiguous villages in an area broadly classified as moderate risk (∼17% PfPR) by recent government stratification [44]. Such fine-scale variability is not uncommon and has been observed in several other settings [75]. In one study in Madagascar, there was a tenfold difference of malaria prevalence within a radius of less than 50 kilometers [76]. For precise micro-stratifications, this study emphasizes the importance of carefully selecting diagnostic tools, especially for local malaria elimination efforts. Our findings indicate that RDTs and microscopy have poor positive predictive values, which can be even less than 20% in villages with very low and low transmission as the proportion of truly infected individuals is very small compared to non-infected persons. There were also significant discrepancies in the resulting micro-strata depending on the test method used. For instance, among the 35 surveyed villages, RDTs and microscopy classified 12 and 11 as very low and 6 and 10 as low risk strata, respectively, while qPCR identified only 1 village as very low and 5 as low transmission. This means RDTs and microscopy classified majority of the villages as very low to low risk while qPCR classified most villages as moderate to high risk (Table 4).

Clear demarcation of areas with very low to low risk versus those with moderate to high risk is essential, particularly in the push towards elimination. As countries increasingly adopt data-driven decision-making for malaria control, there is a risk of improper resource allocation or premature withdrawal of effective interventions from localities erroneously deemed as nearing elimination. Local authorities need to decide which data to use for local-level micro-stratification and whether RDTs, commonly used for broader-scale sub-national stratification, suffice for fine-scale local decision-making. Previous evidence has shown that hotspots identified by RDTs are less stable than those identified by microscopy and PCR [41]. Hotspots of febrile malaria infections are also generally unstable and variable over geographical spaces, while hotspots of asymptomatic cases tend to be more permanent and can be more practically targeted for transmission control [77]. In this current study, we also found significant positive correlations between malaria parasite densities and malaria prevalence in southeastern Tanzania, emphasizing the need to incorporate tests that depict sub-microscopic infections into malaria stratification and decision making to better target the hotspots. As already reported in other studies, villages in our study area, which were classified as low transmission areas had lower geometric mean parasite density compared to those with higher transmission rates.

Our findings, benchmarked against qPCR, reveal limited detection capabilities of RDTs and microscopy in overall fine-scale stratifications, especially in low transmission settings. Previous studies have emphasized the usefulness of routine hospital data for micro-stratifications [8,10,78,79]. However, evidence indicates that both microscopy and RDTs are less effective in identifying stable febrile malaria hotspots, except for asymptomatic hotspots, which are reliably identified by microscopy [77], however still not stable when transmission is low [80]. Our research underscores the importance of identifying subclinical infections using sensitive tools to advance malaria elimination, particularly through fine-scale population surveys. Additionally, there is evidence suggesting potential benefits from integrating hospital and school-age children survey data or even antenatal care centers [2,9]. Nevertheless, these approaches heavily rely on rapid diagnostic tests (RDTs) as the primary tool for malaria detection. While the World Health Organization (WHO) recommends monitoring RDT performance alongside microscopy, our study is particularly relevant as we have directly compared the fine-scale stratification capabilities of RDTs, microscopy, and qPCR at a fine scale.

This study demonstrated overall good agreement between RDTs and qPCR, while microscopy showed fair agreement. However, RDTs missed over 38% of malaria infections, particularly among adults over twenty years old, who were found to harbor lower parasite densities compared to those under twenty years old. However, RDTs remain useful in testing fever-positive malaria cases in hospitals and are widely employed in population surveys due to their cost-effectiveness and ease of implementation [32]. As evidenced in this study, carefully re-consideration of using RDTs for finer-scale mapping and intervention planning at sub-district level should be a priority. Similarly, microscopy missed >50% of the malaria infections detected by qPCR, which is consistent with previous studies, including a meta-analysis of 42 studies, which showed that microscopy misses over 50% of malaria infections [26,66]. Operational challenges, such as the level of expertise required for accurate detection and the need for electricity and precise sample handling procedures, contribute to these limitations. Interestingly, microscopy underestimated malaria risk by classifying more villages as low strata compared to qPCR. Nonetheless, microscopy still plays a crucial role when used in conjunction with tools like RDTs, providing valuable information about malaria parasite densities [81–83]. Here, we estimate a 100-fold higher parasite density when measured by microscopy compared to densities measured by qPCR, consistent with similar trends observed in previous studies [55]. The findings of this study also indicate that the false-negative rate of microscopy decreases with increasing parasite density, a pattern observed in other studies too [81].

The analysis also revealed variations in parasite densities across different age groups, with school-age children (5-15 years old) exhibiting higher parasite densities compared to individuals aged 16 and above. Notably, our study identified a reduced sensitivity of both RDTs and microscopy among adults aged over 16 years, consistent with findings from prior studies conducted before 2015 in various regions [84–91]. It is possible this pattern is driven by the age-related differences in malaria parasite prevalence, which was also observed (Table 3). Furthermore, our findings suggest that this trend may be attributed to the lower parasite density estimates observed in adults within the study (Figure 7A & 7B). Significantly, our research provides valuable insights, highlighting the potential implications of these trends, particularly in fine-scale mapping scenarios, where RDTs and microscopy may underestimate stratifications at very low and low transmission strata, with qPCR serving as the reference standard in this study.

Ultimately, this study underscores the potential need to expand the use of molecular approaches such as qPCR to achieve more precise mapping of malaria infections, which could reduce false negatives and facilitate more precise resource allocations. However, qPCR has operational challenges, including the need for well-designed infrastructure, high costs, and expertise, and it is not portable for remote areas [82]. Efforts are underway to develop portable qPCR technologies, but cost and expertise remain significant barriers. To address these gaps, NMCPs should develop innovative plans, which might include: a) centralized facilities receiving and processing qPCR samples and then doing such surveys only infrequently, say every 3 years; b) partnering with local research organizations to support the high accuracy evaluations with nucleic acid based tests. A related point to emphasize is the overall need for highly-sensitive, cost-effective, and potentially reagent-free tools that align with the economic context of malaria-endemic settings. Recent innovations such as the saliva-based tests [92] or the use of Infrared spectroscopy (IR) and machine learning (ML) [82,93] have shown promise in detecting malaria infections at sensitivities equivalent to PCR, but further research are needed before these technologies can be routinely deployed. Such reagent-free assays like the IR-AI based approaches would be particularly transformative for scaling up effective micro stratification of malaria risk in Africa.

This study also raised some important new questions. For example, it is baffling to observe that areas with low transmission also have persistently low parasite densities despite individuals having low immunity compared to those in higher transmission settings who acquire immunity and become protective. Studies have demonstrated that in low transmission areas, highly virulent parasites are more exposed to facilitate malaria transmission by mosquitoes compared to low virulent ones [26,94–96]. Consequently, high virulent parasites are detected and treated, leading to their removal from the population [26,94–96]. This leaves behind low virulent parasites that are less exposed and maintain low densities, becoming symptomatic, undetectable, and untreated [26,94–96]. This phenomenon may contribute to long-term parasite transmission strategies, highlighting the importance of using highly sensitive tools for screening [55,97,98].

One limitation of this study is its failure to consider factors that may contribute to the broader heterogeneity of malaria infections in southeastern Tanzania. Future investigations should delve into potential environmental, geographical, immunological or genetic diversity of the parasite influences underlying this variability. Additionally, the biological significance of missed infections by both RDTs and microscopy was not explored. Consequently, the study did not estimate the transmission burden associated with these undetected positive samples, nor assess the parasite densities necessary to sustain transmission in the population.

## Conclusion

As countries progress towards malaria elimination, fine-scale mapping of malaria risk becomes increasingly important. This study highlights significant variability in village-level malaria risk within and between districts in southeastern Tanzania, an area where the scale-up of effective interventions has led to substantial progress, yet cases persist despite high intervention coverage. Secondly, the study underscores the variable performance of different testing methods in stratifying risk. While RDTs and microscopy, the primary test methods used in low-income endemic settings and the main sources of data for ongoing epidemiological stratification efforts, were effective in high-transmission areas, they performed poorly in low-transmission settings, often classifying most villages as very low or low risk. In contrast, qPCR classified most villages as moderate or high risk. These findings demonstrate the importance of using appropriate testing methods for data-driven, fine-scale risk stratification to enhance targeted interventions aimed at reducing and eliminating malaria. The study underscores the need for improved testing approaches that are both operationally feasible and sufficiently sensitive to enable precise mapping and effective targeting of malaria in local contexts. More importantly, public health authorities must recognize the strengths and limitations of their available data when planning local stratification or making decisions. While innovation for more effective strategies is ongoing, sensitive molecular tools like qPCR, despite their operational challenges, will be crucial for accurate malaria risk mapping and intervention planning, especially in settings with significantly reduced risk. Going forward, developing new tools that balance operational costs and sensitivity, particularly in low transmission settings, will be essential for effective malaria control and eventual elimination.

## Acknowledgements

We extend our sincere gratitude to the dedicated team of the Deep Diagnostics Project at Ifakara Health Institute for their invaluable support throughout this survey. Special recognition is due to Abigael W. Magesa, Tumpe G. Mwandyala, Magreth C. Magoda, Scolastica N. Hangu, Alex T. Ngonyani, Magreth I. Henry, Jiddah A. Ally, Alfred J. Simfukwe, Amisa N. Rajabu, Scolastica A. Ndonde, Rahma S. Kikweo, Benson D. Makua, Jacob J. Nyooka, Festo I. Tangaliola, Stephen J. Magasa, Joyce Y. Jackson, Fidelis D. Mbena, Slyakus V, Mlembe, and Agnes Kimaro for their invaluable contributions to conducting the surveys, administering questionnaires, screenings, and laboratory procedures. We also express our gratitude to Judith Mshumbusi and the procurement team for their assistance in procuring materials, often on short notice. Additionally, we extend our thanks to Faraji Abilahi, the Ifakara laboratory manager, and fellow scientists, students and staff at Ifakara Health institute and University of Glasgow, who contributed direct or indirectly to this study.

## Author contributions

Conceptualization: I.H.M., F.O., F.B., and S.A.B. Data curation: I.H.M., F.M., E.G.M., I.M., and F.E.M. Formal analysis: I.H.M., S.A.B., F.O.O., D.B.,H.S.N., F.B. Funding acquisition: F.O.O, S.A.B., F.B., and I.H.M. Investigation: I.H.M., F.M., E.G.M., I.M., N.S.L, S.A, and F.E.M. Methodology: I.H.M., A.B.L., S.A.B., F.O.O., F.B. Project administration: I.H.M., R.M.N., F.M., Resources: I.H.M., F.M.,H.S.N S.A.B., F.O.O., and F.B. Supervision: S.A.B., F.O.O., and F.B. Validation: S.A.B., F.O.O., and F.B. Visualization: I.H.M., S.A.B., F.O.O., F.B., N.F.K, A.L. D.B. Writing original draft: I.H.M., S.A.B., F.O.O., and F.B. Writing review & editing: All authors.

## Ethics approvals

This work was approved under Ifakara Health Institute Review Board (Ref: IHI/IRB/No: 1 2021) and the National Institute for Medical Research-NIMR (NIMR/HQ/R.8a/Vol. 1X/3735). Permission to publish this work has been granted with reference No: BD. 242/437/01C/6.

## Data availability

The dataset supporting the findings is available upon a reasonable request, which can be directed to either the corresponding author, the Ifakara Health Institute ethical review board in Tanzania or National Health Research Ethics Committee in Tanzania with reference to ethical clearance certificate of NIMR/HQ/R.8a/Vol. 1X/3735.

