## Supplementary Table 1 and 2 for "Comparison of Fine-Scale Malaria Strata Derived from Population Survey Data Collected Using mRDTs, Microscopy and qPCR in South-Eastern Tanzania"

**Table 1:** Preparation of 1 mL of 5x Oligo mix and their respective sequence

| **Oligo name** | **Species specificity** | **Target region** | **Oligo sequence** | **Oligo modification [5’-3’]** |
| --- | --- | --- | --- | --- |
| ***P. falciparum* (PlasQ assay)** |  |  |  |  |
| Pspp18S F | *Plasmodium spp* | 18S rDNA | GCT CTT TCT TGA TTT CTT GGA TG | - |
| Pspp18S R | *Plasmodium spp* | 18S rDNA | AGC AGG TTA AGA TCT CG TTC G | - |
| Pspp18S probe | *Plasmodium spp* | 18S rDNA | ATG GCC GTT TTT AGT TCG TG | Cy5-BHQ2 |
| PfvarATS F | *P. falciparum* | varATS | CCC ATA CAC AAC CAA YTG GA | - |
| PfvarATS R | *P. falciparum* | varATS | TTC GCA CAT ATC TCT ATG TCT ATC T | - |
| PfvarATS probe | *P. falciparum* | varATS | TRT TCC ATA AAT GGT | FAM-NFQ/MGB |
| HsRNaseP F | *H. sapiens* | RnaseP gene | AGA TTT GGA CCT GCG AGC G | - |
| HsRNaseP R | *H. sapiens* | RnaseP gene | GAG CGG CTG TCT CCA CAA GT | - |
| HsRNaseP probe | *H. sapiens* | RnaseP gene | TTC TGA CCT GAA GGC TCT GCG CG | YakimaYellow-BHQ1 |

**Table 2:** Master Mix preparation

| **Component** | **Stock concentration** | **Final concentration** | **Reaction volume (μL)** | **Example for 100 reactions (μL)** |
| --- | --- | --- | --- | --- |
| Luna Universal Probe qPCR Master Mix | 2x | 1x | 5 | 500 |
| PlasQ Primer Mix | 5x | 1 | 2 | 200 |
| Molecular biology grade H2O | - | - | 1 | 100 |
